# Sparse canonical correlation to identify breast cancer related genes regulated by copy number aberrations

**DOI:** 10.1101/2021.08.29.21262811

**Authors:** Diptavo Dutta, Ananda Sen, Jaya Satagopan

## Abstract

**Background:** Copy number aberrations (CNAs) in cancer affect disease outcomes by regulating molecular phenotypes, such as gene expressions, that drive important biological processes. To gain comprehensive insights into molecular biomarkers for cancer, it is critical to identify key groups of CNAs, the associated gene networks, regulatory modules, and their downstream effect on outcomes.

**Methods:** In this paper, we demonstrate an innovative use of sparse canonical correlation analysis (sCCA) to effectively identify the ensemble of CNAs, gene networks and regulatory modules in the context of binary and censored disease endpoints. Our approach detects potentially orthogonal gene expression modules which are highly correlated with sets of CNA and then identifies the genes within these modules that are associated with the outcome.

**Results:** Analyzing clinical and genomic data on 1,904 breast cancer patients from the METABRIC study, we found 14 gene modules to be regulated by groups of proximally located CNA sites. We validated this finding using an independent set of 1,077 breast invasive carcinoma samples from The Cancer Genome Atlas (TCGA). Our analysis on 7 clinical endpoints identified several novel and interpretable regulatory associations, highlighting the role of CNAs in key biological pathways and processes for breast cancer. Genes significantly associated with the outcomes were enriched for early estrogen response pathway, DNA repair pathways as well as targets of transcription factors such as E2F4, MYC and ETS1 that have recognized roles in tumor characteristics and survival. Subsequent meta-analysis across the endpoints further identified several genes through aggregation of weaker associations.

**Conclusions:** Our findings suggest that sCCA analysis can aggregate weaker associations to identify interpretable and important genes, networks and pathways that are clinically consequential.

## Introduction

Cancer genomes have an enriched burden of somatic copy number aberrations^1–3^ (CNAs) such as DNA copy number gains and losses that harbor or are proximal to important oncogenes and tumor suppressor genes controlling cell growth and division^4^. The CNAs may directly regulate cellular growth pathways and other gene sets impacting key biological outcomes by altering gene expressions at the RNA level that are important for tumorigenesis and outcomes. Thus, identification of such driver CNAs and understanding the mechanism through which they affect downstream gene expression and the resulting effects on cancer outcomes are crucial for effective disease management and control.

This past decade has witnessed significant advances in our ability to measure large volumes of data on CNA and gene expressions from tumor samples. These data offer unprecedented opportunities to identify biomarkers associated with disease outcomes. These opportunities have accelerated a shift towards the development of novel genomics-based prognostic and therapeutic tools. For example, CNAs are associated with poor survival in breast cancer patients^5^; mutations in *PIK3CA* are associated with poor survival in certain estrogen receptor (ER)-positive breast cancers^6,7^. Oncotype DX, the FDA-approved score based on the expression of 21 genes, and MammaPrint, a score based on the expression of 70 genes, are associated with poor survival in breast cancer patients and are used for making treatment decisions in estrogen-positive early-stage breast cancers and all early-stage breast cancers, respectively^8–11^. Identifying such biomarkers have been pivotal to advancing cancer care in the past decade. However, due to low prevalence of specific CNAs and since existing gene expression scores are relevant for only specific subgroups of cancer patients, current biomarkers can only be used for managing the disease of a small segment of patients. Given the growth and aging of the US population and, with this, the projected 50% increase in cancer incidence over the next three decades^12^, further research is urgently needed to identify biomarkers for effective management of all types of cancers. In this article we present a novel statistical and computational approach to address this need, focusing on CNAs and gene expressions in breast cancer.

Standard methods for identifying biomarkers examine the association between individual genomic features and outcome (such as survival) and select the top-ranking biomarkers after adjusting for multiple comparisons^13–15^. Penalized regression methods^16^, including the causal modeling with expression linkage for complex traits (Camelot) approach^17^, relate multiple CNAs and gene expressions to an outcome via a multivariable regression model to select biomarkers by imposing sparsity via L_1_ or L_2_ penalties. These methods prioritize biomarkers based on their individual effects on the outcome without leveraging the putative biological relationships between CNAs and gene expressions. It is well-understood that genes and their products rarely act in isolation but work with other genes or their products to form networks or pathways to address specific biological functions^18^. Thus, a comprehensive understanding of gene networks at DNA and RNA levels and the resulting impact on disease outcomes is warranted for identifying clinically relevant biomarkers. To this end, the piecewise linear regression spline (PLRS) method^19^, the lots of lasso (Lol) method^20–22^, the weighted correlation network analysis^23^, and Oncodrive-CIS^24^ identify significant CNA-gene expression pairs, but do not provide comprehensive maps of regulatory networks that are essential for gaining insights into biomarkers reflecting disease biology. The relationship between networks of CNAs and downstream networks of gene expressions and the resulting effect on cancer outcomes have not been fully explored.

The key challenges in analyzing such networks are the large dimensionality of the CNA and gene expression data sets and the sparsity of CNAs. The latter issue leads to considerable challenges for modeling large and sparse matrix of CNAs in relation to large gene expression matrix to identify combinations or networks of somatic changes that regulate combinations or modules of gene expressions. A pragmatic strategy to overcome this challenge would be to view this as a sparse canonical correlation analysis (sCCA)^25,26^ problem to produce sparse latent variables representing biologically relevant CNA sets and gene expression modules or networks to achieve maximal correlation between the CNA and gene expression matrices and relate the resulting gene expression modules to disease outcomes to identify the biomarkers of interest. This paper is based on the thesis that substantial biological and clinical relevance for cancer outcomes can be effectively captured using a sparse or smaller number of CNA sets and gene expression networks. But this requires innovative statistical and computational application of sCCA to extract such sparse latent variables effectively.

We present a two-step analysis framework that aims to map networks of CNAs that regulate modules of gene expressions to affect cancer related outcomes. In the first step, sCCA is used to identify gene expression modules that are regulated by CNA networks in an unsupervised manner that does not employ the disease outcomes. Given the gene modules, in the second step we use a multivariable generalized linear model framework to isolate genes within a particular gene expression module that are associated with breast cancer related outcomes. The key advantage of our proposed approach is that it is particularly amenable to interpretation as it not only identifies the genes whose expression levels are associated with outcomes but also identifies the set of CNA which potentially regulates them.

We apply our approach to analyze data on 1904 breast cancer patients from the METABRIC study^7,27^ for whom data are available on CNAs, gene-expressions, and various clinical information related to breast cancer (See Supplementary Methods for details on the study). As an example analysis, we focus on two clinical outcomes: estrogen receptor status, which is a binary variable, and overall survival, which is a censored variable measured as months elapsed from the date of study entry to date of death or, if the patient is alive, the end of the study. Through extensive downstream analysis we demonstrate that the genes identified to be associated to the clinical outcomes have plausible independent evidence to be biologically relevant for the outcome of interest. Further, we meta-analyzed the results across six different clinical outcomes to identify genes that were potentially associated with multiple outcomes. We additionally validated the gene modules identified from the METABRIC study in an independent data on individuals in a study of breast invasive carcinoma from The Cancer Genome Atlas (TCGA) that includes CNAs, gene expressions, clinical variables, and outcomes in 1,077 patients. Both the data sets were obtained from the cBioPortal catalog^28^.

## Overview of methods

To describe our approach, we assume that we have individual level data for n individuals on p copy number aberrations (CNA) and q gene-expressions.

### Step 1. Identifying Gene modules through sCCA

We first aim to identify gene modules regulated by CNA, by mapping groups of CNA to groups of associated gene expressions using sparse canonical correlation analysis (sCCA). sCCA identifies approximate orthogonal gene modules that are regulated by CNA. This step is agnostic of any phenotypic information or outcomes. For *n* individuals, let ***G***^***n*×*p***^ be the matrix for *p* sites of copy number aberration (CNA) sites with ***Gij*** being the number of insertion or deletions for individual i at site j, and ***E***^***n*×*q***^ be the normalized gene-expression levels for *q* genes across n individuals. Sparse canonical correlation analysis (sCCA) identifies sparse linear combinations of CNA (***u***^***p*×1**^; termed CNA component) and gene-expressions (***v***^***g*×1**^; termed gene component) such that the correlation between ***Gu*** and ***Ev*** is maximized i.e.,

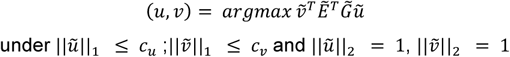

where ||·||_***h***_ denotes the *L*_*h*_ norm and 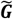 (or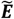)denotes the normalized version of the corresponding matrix. The subsequent pairs of sCCA components are obtained similarly by matrix deflation and under the constraint of being uncorrelated or orthogonal to the previous components. Ideally each pair of sCCA components selects a sparse set of CNA sites that regulate the expression of a sparse set of genes across the genome, denoted by the non-zero elements in *u* and *v* respectively. Overall, the sCCA aggregates multiple associations between the selected CNA and genes and hence represents principal regulation or association patterns. Additionally, due to the orthogonality constraint each pair of sCCA component reflect approximately an independent or orthogonal pattern of regulation. *cu* (and *cv*) represent the sparsity parameters for the CNA and gene components respectively. To facilitate interpretation, we choose the sparsity parameters such that there is no overlap between the CNA selected in the components. (See Supplementary Methods for details).

### Step 2. Association with outcomes

Given the gene-modules identified in Step 1, we now identify which genes within these modules are associated with the outcomes of interest.

#### Univariate outcomes

Let 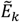 be the *n* **×** *rk* matrix of normalized gene-expressions for the genes selected in sCCA component *k*, where *rk* = ||*vk*||0 and *vk* denotes the gene-component of the kth sCCA component. We use the following generalized linear model to associate the *rk* genes to a phenotype *y* as

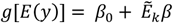

Where *g*[·] is a canonical link function and *β, β*_0_ are regression parameters. For each of the gene modules identified by sCCA, we perform the association analysis and record gene-specific p-values and obtain the false discovery rates (FDR). Genes with FDR < 0.05 are declared to be significantly associated with the outcome.

#### Multivariate outcomes

If multiple, potentially correlated, outcomes are available for the individuals, we can meta-analyze results across the multiple outcomes to identify genes that are possibly associated to more than one outcome. Let *p*_1_, *p*_2_, …,*ps* be the univariate p-values for a particular gene for s outcomes, from the previous univariate association analysis. These p-values are likely to be correlated due to potential correlation between the outcomes. We perform a cauchy-transformed meta-analysis^29^ which has been shown to maintain correct false positive rate in presence of correlation as well^30,31^. Specifically, we transform each of the p-values to a cauchy variable as

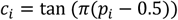

The test statistic is the unweighted mean of these transformed variables which follows a standard cauchy distribution, under the null hypothesis of no association, irrespective of the correlation between the outcomes^29^.

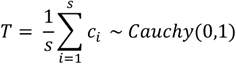

The overall p-value can be calculated by inverting the cumulative density function of the standard cauchy distribution.

## Results

We started with 1,904 individuals who had complete data at 22,544 CNA sites and expression level data for 24,360 genes. Sparse canonical correlation analysis (See Methods) identified 14 gene modules through the sCCA components. In this article, we will use the terms *modules* and *networks* synonymously to denote the collection of genes selected in a gene component and *set* to denote the CNA sites selected in a CNA component. Across the 14 gene modules, sCCA selects 824 genes, whose expression levels are regulated by 1,851 CNA sites overall (Table 1). In general, for each sCCA component, the CNA component selects CNA sites located in a small sub-region within a chromosome (Figure 1A). Our sCCA analysis was agnostic of the physical location of the CNA in the genome and hence the sCCA algorithm is not guided or biased towards selecting positionally proximal CNA. However, due to the high correlation between nearby CNA, each CNA component selects a smaller subregion in chromosome of high correlated CNA which might have regulatory effects on the gene selected in the corresponding gene component. For example, the 115 CNA sites selected in CNA component 4, were located on chromosome 17q11.2-q21.32 region. The corresponding gene components can then be viewed as the gene module (or network) having strong association to or being potentially regulated by the selected CNA sites and can possibly mediate their effects. In general, we expect the regulatory structures captured by the sCCA components to be approximately independent. However, we notice that the expression levels of genes selected in gene component 8 has a higher correlation with the CNA selected in CNA component 2 (Figure 1B). It is to be noted that CNA components 2 & 8 defined highly proximal regions in chromosome 8. Hence, the correlation between gene module 8 & CNA set 2 is not unexpected due to LD and/or possible long range regulatory activity. This indicates that genes modules 2 and 8 are possibly coregulated by the CNA selected in the corresponding CNA components. Overall, a CNA component defines a chromosomal subregion which has potentially multiple independent regulatory effects on the gene module identified by the corresponding gene component. The advantage of the sCCA in this application is that it can aggregate multiple, possibly weaker association to select groups of CNA associated with genes modules (See Supplementary Table 1-2 for full list of CNA and genes selected).

**Table 1:** Description of the 14 gene modules and CNA components identified through sCCA.

|  | CNA components |  |  | Gene components (modules) |  |  |
| --- | --- | --- | --- | --- | --- | --- |
| sCCA component | Number of CNA selected | Chromosome Number | Genomic location of selected CNA (Mb) | Number of Genes selected | Genes on different chromosome | Distal genes on same chromosome |
| 1 | 139 | 5 | 72.17 – 119.54 | 70 | 24 | 18 |
| 2 | 107 | 8 | 135.45 – 145.01 | 59 | 0 | 14 |
| 3 | 131 | 1 | 153.20-156.96 | 56 | 0 | 6 |
| 4 | 115 | 17 | 41.86 – 45.50 | 67 | 9 | 4 |
| 5 | 145 | 22 | 35.64 – 44.57 | 63 | 0 | 6 |
| 6 | 130 | 16 | 65.28 – 77.25 | 58 | 0 | 6 |
| 7 | 125 | 22 | 17.95 – 24.63 | 58 | 0 | 10 |
| 8 | 125 | 8 | 115.95 – 134.48 | 59 | 0 | 24 |
| 9 | 145 | 7 | 100.40 – 127.61 | 71 | 0 | 17 |
| 10 | 151 | 9 | 121.82 – 131.49 | 74 | 0 | 1 |
| 11 | 129 | 16 | 1.06 – 3.72 | 64 | 1 | 11 |
| 12 | 117 | 1 | 9.65 – 16.64 | 63 | 0 | 12 |
| 13 | 147 | 3 | 168.00 – 193.44 | 71 | 0 | 11 |
| 14 | 145 | 17 | 58.16 – 69.14 | 62 | 0 | 1 |

**Figure 1:**
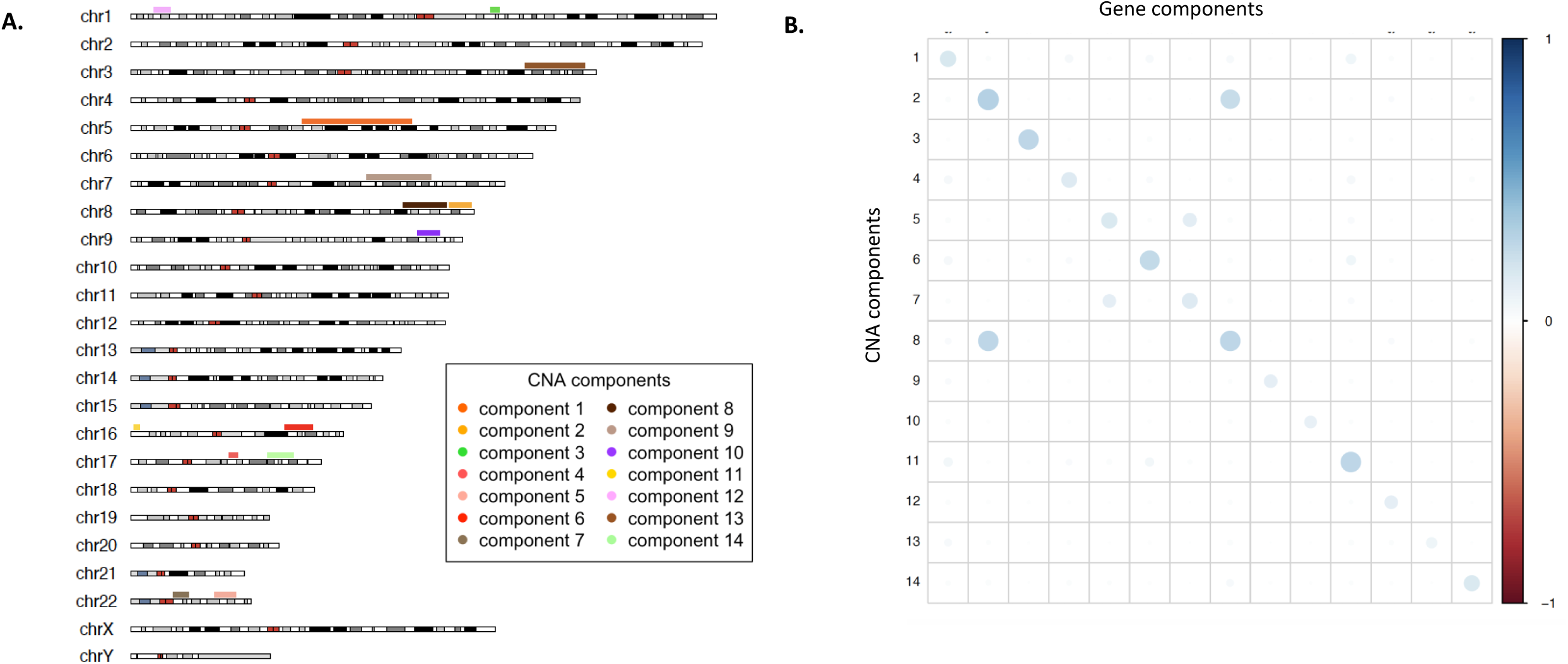
Results from sCCA analysis. (A) Chromosomal subregions identified by 14 CNA components. For each CNA component the region between the most distal CNA selected in that component is marked. (B) Average squared correlation between the CNA selected in CNA components and genes selected in Gene components. Correlation between Genes in components 2 and CNA in component 8 (and vice versa) are observed due possible correlation between selected CNA and long-range regulatory effects. Similar correlation is observed for components 5 & 7 as well.

### Gene modules capture cis and trans effects

Through the identification of gene modules, we capture regulatory effects of CNA. In general, we found that most of the associations aggregated by the sCCA components identified effects of CNA sites on nearby (cis) gene expression. On average, 44.8% of the genes selected in each sCCA component also has a CNA in or near the same gene selected in the respective CNA component. This is expected since cis effects are known to be much stronger compared to distal (trans) effects and would have a direct regulatory effect on the expression level of a nearby gene. However, several examples of distal (trans) regulatory effects on expressions of genes on different chromosome were also identified in the gene modules (Figure 2A-B). On average, 3.2% of the genes selected in the gene components were on a different chromosome than the corresponding CNA component. Further, on average 15.9% of the genes selected in the gene components were more that 10Mb away from the sub-region of chromosome selected by the corresponding CNA component, indicating long range regulatory effects (Table 1). For example, among the 67 genes selected in component 4, 9 genes are on different chromosomes and an additional 4 genes are outside of the region 17q11.2-q21.32, which contains the CNA selected by CNA component 4. We found possible mechanistic explanations for several such distal associations in existing genomic and profiling data. For example, gene component 4, selects atlastin GTPase 3 gene (*ATL3*) on chromosome 11. *ATL3* is a downstream target for transcription factor Signal Transducer and Activator of Transcription 3 (*STAT3*) in ENCODE transcription factor database^32,33^. Interestingly, a copy number aberration of *STAT3* was selected among the CNA sites in CNA component 4, which suggests a possible cis-mediation mechanism for the association of this and other nearby CNA sites with *ATL3*.

**Figure 2:**
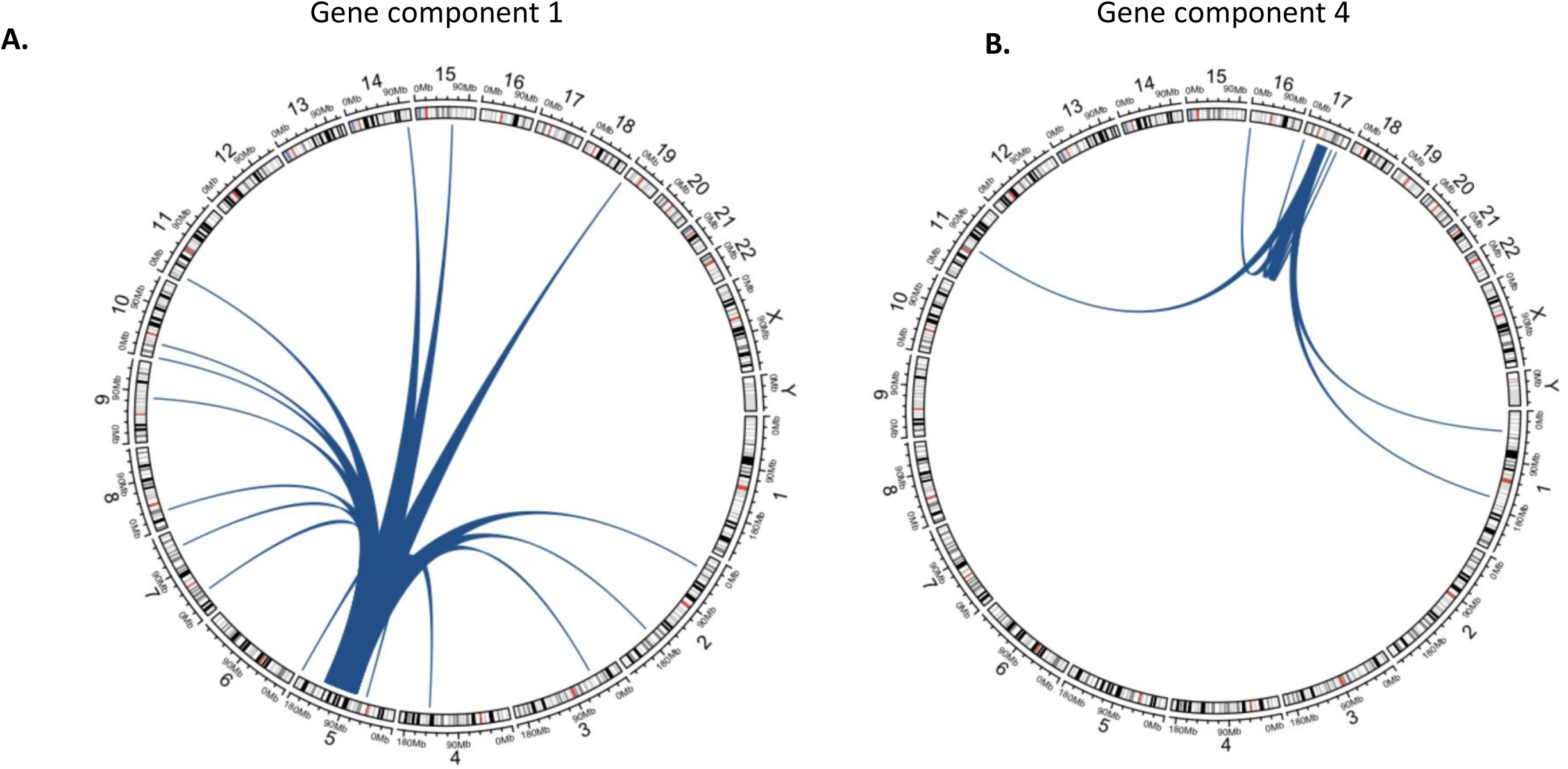
Examples of *trans*-associations. identified in using the genes selected through sCCA in (A) Gene component 1 (B) Gene component 4. Several genes in a chromosome different from that of the selected CNA is identified. Further, numerous distal genes (> 10Mb) on the same chromosome are detected as well.

### Evidence of coregulation

To further validate whether the genes selected by the 14 significant sCCA components had any overall evidence of biological coregulation as well, we used large-scale transcription factor databases from the ENCODE study^32^ and existing ChIP-chip, ChIP-seq, and several other transcription factor binding site profiling experiments (ChEA)^34,35^. For the 181 transcription factors and their downstream targets reported in ENCODE, we found that, across the 14 gene module identified though sCCA components, on an average 67.3% of the genes were downstream targets for more than 20 transcription factors. For ChEA, which reports data on 202 TFs and their downstream targets, we found similarly that on average 65.1% genes were downstream targets for more than 20 transcription factors. This provides implicit evidence that a large proportion of the genes selected by the sCCA components might have evidence of being coregulated by TFs and the identification of gene modules using sCCA analysis can successfully detect such patterns of coregulation as an independent line of evidence.

### Replication of Gene Modules using TCGA breast invasive carcinoma data

Selection of genes and CNA can be influenced and biased if there are systematic biases and batch effects. So, we investigated whether the gene modules and CNA sites identified through sCCA, were replicable in an external dataset. For that, we used the TCGA breast invasive carcinoma data (See Supplementary Methods for details on the study), which reports data on CNA sites and gene expression in primary breast tumor tissue for 982 breast cancer patients. We adopted a resampling-based procedure to test whether the sCCA components represented gene modules and CNA sets that had stronger association than expected at random. For a given gene module (selected through a gene component), we evaluated whether the observed average squared correlation between these genes and CNA selected in corresponding CNA component were higher than what is expected at random. Similarly, for a set of CNA (selected through a CNA component), we evaluated whether the observed average squared correlation between these CNA and genes selected in corresponding gene component were higher than what is expected at random. We found that among the gene modules and CNA sets selected in METABRIC and present in TCGA, the average correlation for all the 14 components were significantly (p-value < 0.05) higher than expected (Supplementary Figure S1). Further 10 of these components were strongly significant as well (p-value < 0.001). Such a result is not unexpected as the sCCA components include a majority of stronger cis effects. Further, this also suggests that the sCCA components in METABRIC possibly captured true effects replicable across different datasets and not potential artefacts and batch effects within METABRIC. (See Supplementary Methods)

### Association with breast cancer related outcomes

Given the 14 gene modules obtained through sCCA analysis, we investigated whether these gene modules were associated with 7 different types of breast cancer outcomes (Table 2). At a lenient cutoff of FDR < 0.05, we found that 562 genes across the 14 modules were associated with at least one of the outcomes (Supplementary Table 3). Further, at a stringent exome-wide cutoff of p-value < 2.5 × 10^−06^, we found 94 genes associated with at least 1 outcome. Subsequently, through several downstream analysis we investigated whether the genes that are significant for a given outcome indeed had external evidence of association to breast cancer related outcomes. Here we demonstrate the results for two distinct types of outcomes:

**Table 2:** Description of the seven breast cancer related outcomes analyzed, and the number of genes associated significantly.

| Outcome | % cases | Median survival | Significant Genes (FDR < 0.05) |
| --- | --- | --- | --- |
| Estrogen Receptor status (ER) | 76.6 | - | 210 |
| Progesterone receptor status (PR) | 52.9 | - | 237 |
| Human Epidermal growth factor Receptor 2 status (HER2) | 12.4 | - | 255 |
| Grade (Grade 3 vs Grade lower than 3) | 47.5 | - | 65 |
| Lymph Nodes Examined to be present (present vs absent) | 47.8 | - | 12 |
| Age at diagnosis (< 50 years) | 78.4 | - | 100 |
| Overall Survival | - | 154.2 | 73 |

### Estrogen Receptor (ER)

Of the 1,904 individuals in the sample, 1,459 (76.6%) individuals had ER positive status. We performed logistic regression-based association tests of the 14 significant gene modules. Across the components we found that 210 genes were significant at an FDR < 0.05 and 36 genes were significant with p-value < 2.5 × 10^−06^. Among the genes significantly associated with ER status, we identified known breast cancer related genes such as Microtubule Associated Protein Tau (*MAPT*), whose expression is highly associated with low sensitivity to taxanes that are important drugs for breast cancer treatment^36^, and Macrophage migration inhibitory factor (*MIF*), a pro-inflammatory cytokine whose blockade reduces the aggressiveness of invasive breast cancer^37^. Further our sCCA-based model provides a potential explanation of the related biological mechanism. For example, among genes selected in gene component 4, we found that Dynein Axonemal Light Intermediate Chain 1 (*DNALI1*), on chromosome 1 is associated with ER status (p-value = 4.8 × 10^−04^), being trans-regulated by CNA sites on chromosome 17 selected in CNA component 4 (Figure 3A). *DNALI1* is a downstream target for transcription factors *STAT3* and *UBTF*, both of which are selected in CNA component 4. Further, there is evidence of physical interactions between the proteins resulting from *DNALI1* and *UBTF* in large protein interactions databases as well^38^. This indicates the possibility that *DNALI1* mediates the effects of the CNA sites in chromosome 5 selected by CNA component 1, on ER status. Thus, not only we identify the genes whose expression levels are associated with breast cancer outcomes, we also additionally identify which CNA potentially regulate such genes.

**Figure 3:**
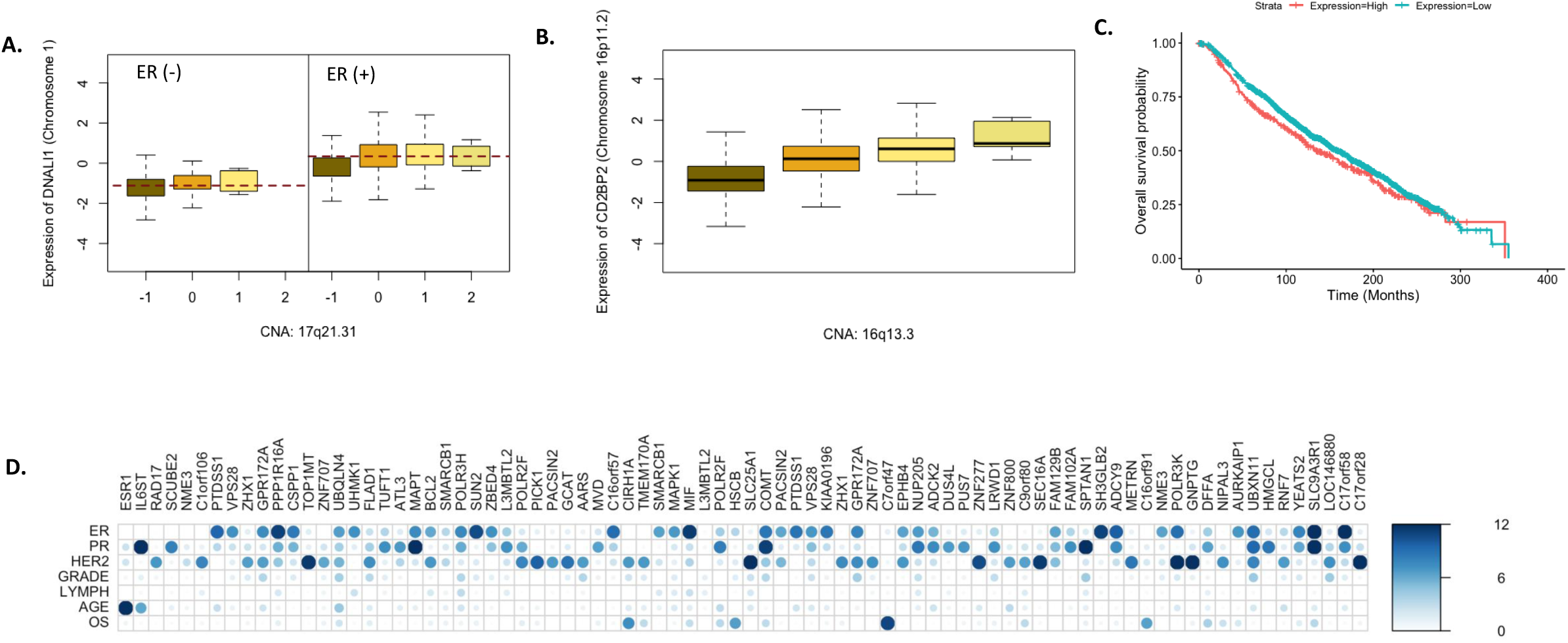
Association analysis with breast cancer related outcomes. (A) *DNALI1* gene on chromosome 1 associated with ER status and trans-regulated by a CNA of transcription factor *CHD1* on chromosome 5. (B) Trans regulation of *CD2BP2* gene on chromosome 16p11.2 by a CNA in *ZNF263* gene located in chromosome 16q13.3 which are approximately 27Mb apart. (C) Association of *CD2BP2* expression level with overall survival probability. Expression levels have been dichotomized as high and low using 75-th percentile as cut-off. (D) p-values of 72 genes identified to be strongly associated (p-value < 2.5 × 10^−06^) with multiple outcomes, across the 7 outcomes. P-values < 1 × 10^−12^ are collapsed to 1 × 10^−12^ for the ease of viewing.

Through pathway enrichment analysis (Table 3), we found that the genes significantly associated to ER status at FDR < 0.05, were enriched for hallmark pathways^39^ like early response to estrogen, DNA repair and *MYC* targets, *MYC* being a well-known oncogene^40^. Further, in pathways curated from chemical and genetic perturbation experiments, we found that the genes were enriched for genes highly positively co-expressed with BRCA1 and BRCA2, two genes well reported to be involved in breast cancer^41–43^. Further, the genes were also enriched for targets of several transcription factors, like *NELFE*^44,45^, *E2F4*^46,47^ and *CREB1*^48,49^ which are known to play key roles in several cancers, including breast cancer. However, overlap of the identified significant genes with key cancer related pathways suggest a possible mechanistic explanation for the outcome. For example, of the 210 significant genes, 7 genes (*ADCY9, ABAT, MAPT, SLC9A3R1, CANT1, BCL2, FAM102A*) are in the early estrogen response hallmark pathway^39^. These 7 genes are identified as part of gene modules 4, 11, 14, 7 and This indicates that the CNA selected in the corresponding CNA components, which regulates these gene components respectively, as shown in sCCA analysis, significantly changes estrogen response and can possibly be causal for ER status. So essentially perturbations in early estrogen response hallmark pathway can occur due to CNA in chromosomal sub-regions defined by components 4, 11, 7 and 14. This interpretability is a key advantage of our analysis approach.

**Table 3:** Different categories of pathways enriched for the 210 genes associated (FDR < 0.05) with ER status.

| Category | Pathway | Adjusted p-value | Genes in pathway | Genes overlap |
| --- | --- | --- | --- | --- |
| GO | Cellular macromolecule localization | $5.7 \times 10^{-09}$ | 1886 | 39 |
| | Intracellular protein transport | $1.2 \times 10^{-07}$ | 1156 | 28 |
| | Cellular response to DNA damage stimulus | $4.4 \times 10^{-07}$ | 841 | 23 |
| | Catalytic complex | $4.7 \times 10^{-10}$ | 1552 | 36 |
| | Adenyl Nucleotide binding | $3.9 \times 10^{-06}$ | 1536 | 30 |
| Hallmark | Estrogen Response (early) | $4.9 \times 10^{-03}$ | 200 | 7 |
| | DNA repair | $4.9 \times 10^{-03}$ | 149 | 6 |
| | E2F targets | $9.3 \times 10^{-03}$ | 200 | 6 |
| | MYC targets | $9.3 \times 10^{-03}$ | 200 | 6 |
| | MTORC1 signaling | $4.5 \times 10^{-02}$ | 200 | 5 |
| Curated Gene sets | NIKOLSKY_BREAST_CANCER_8Q23_Q24_AMPLICON | $8.2 \times 10^{-14}$ | 157 | 17 |
| | PUJANA_BRCA1_PCC_NETWORK | $2.3 \times 10^{-08}$ | 1617 | 34 |
| | CHARAFE_BREAST_CANCER_LUMINAL_VS_MESENCHYMAL_UP | $4.3 \times 10^{-07}$ | 446 | 17 |
| | VANTVEER_BREAST_CANCER_ESR1_UP | $5.7 \times 10^{-05}$ | 208 | 15 |
| | PUJANA_BRCA2_PCC_NETWORK | $5.4 \times 10^{-04}$ | 423 | 12 |
| Immunologic Signatures | GSE4984_GALECTIN1_VS_VEHICLE_CTRL_TREATED_DC_DN | $1.4 \times 10^{-06}$ | 198 | 12 |
| | GSE2770_UNTREATED_VS_IL12_TREATED_ACT_CD4_TCELL_2H_DN | $3.8 \times 10^{-06}$ | 200 | 12 |
| | GSE19825_NAIVE_VS_IL2RAHIGH_DAY3_EFF_CD8_TCELL_DN | $1.8 \times 10^{-05}$ | 200 | 11 |
| TF targets | ENCODE: NELFE | $6.5 \times 10^{-46}$ | 9442 | 173 |
| | ENCODE: E2F4 | $2.8 \times 10^{-34}$ | 12626 | 180 |
| | ENCODE: CREB1 | $5.7 \times 10^{-33}$ | 12289 | 177 |
| | ChEA: EGR1 | $3.1 \times 10^{-09}$ | 5000 | 82 |
| | ChEA: ELF3 | $2.2 \times 10^{-07}$ | 1760 | 40 |

We further benchmarked whether the 210 genes found to be significant using this sCCA based approach against 161 gene selected through a standard sparse logistic regression (See Supplementary Methods). Assessing the model fit for these two approaches in TCGA data, we found that the BIC of the model including genes significant through the sCCA-based analysis was substantially lower than the model that included the genes selected through sparse logistic regression. This indicates that our approach not only provides a better interpretability of the overall genetic mechanism but also produces comparable or better model fit for the data.

### Overall survival (OS)

Of the 1,904 individuals in the sample, 1,109 (76.6%) individuals died during the study, with median survival time being 154 months approximately. In a cox proportional hazard model, we found 73 genes to be significant (FDR < 0.05) across the 14 components. Notably, several interesting distally regulated genes are identified in our analysis. For example, in sCCA component 11, we found that the expression of *CD2BP2* gene on chromosome 5 is associated with the overall survival. This gene is differentially regulated in T47D cells of breast cancer patients in response to tamoxifen^50^, a widely used hormonal therapy drug for breast cancer^51^. The transcription start site for *CD2BP2* is over 27 Mb downstream from the subregion of chromosome 5 selected through the CNA component The corresponding selected set of CNA in component 11 contains a TF *ZNF263*, which has a long-range regulatory effect on *CD2BP2* on the same chromosome^32^ and indicates that possibly *CD2BP2* mediates the effects of the selected CNA producing significant change in overall survival probability (Figure 3B-C).

A comprehensive pathway enrichment analysis (Table 4) reveals that the selected genes are enriched in gene-sets and pathways defined by several breast cancer related perturbation experiments. For example, we found a significant enrichment of the genes associated to OS, in the genes related to adipogenesis. Enrichment was found among genes up regulated in early primary breast tumors expressing ESR1 vs the ESR1 negative ones. In addition, the genes significantly associated to OS were enriched for targets of several key cancer TFs like *ELK1, YY1* and *RUNX3*^52–54^. As highlighted above, through the sCCA components and the subsequent cox PH association model, we not only identify which genes are associated with OS but also detect the CNA sites regulating these gene expressions and hence affecting OS.

**Table 4:** Different categories of pathways enriched for the 73 genes associated (FDR < 0.05) with overall survival (OS)

| Category | Pathway | Adjusted p-value | Genes in pathway | Genes overlap |
| --- | --- | --- | --- | --- |
| GO | RNA metabolic process | $4.2 \times 10^{-03}$ | 1542 | 14 |
| | Macromolecule catabolic process | $4.2 \times 10^{-03}$ | 1366 | 13 |
| | Cellular component disassembly | $1.2 \times 10^{-02}$ | 537 | 8 |
| Hallmark | Adipogenesis | $2.8 \times 10^{-02}$ | 200 | 4 |
| Curated Gene sets | DIAZ_CHRONIC_MEYLOGENOUS_LEUKEMIA_UP | $1.2 \times 10^{-03}$ | 1397 | 14 |
| | Reactome: Metabolism of RNA | $1.3 \times 10^{-03}$ | 668 | 10 |
| | NIKOLSKY_BREAST_CANCER_17Q21_Q25_AMPLICON | $2.1 \times 10^{-02}$ | 332 | 6 |
| Immunologic Signatures | GSE3982_NEUTROPHIL_VS_TH1_DN | $4.8 \times 10^{-04}$ | 199 | 7 |
| | GSE3982_EOSINOPHIL_VS_NKCELL_DN | $1.5 \times 10^{-03}$ | 197 | 5 |
| | GSE27786_NKCELL_VS_NEUTROPHIL_UP | $8.1 \times 10^{-03}$ | 199 | 5 |
| TF targets | ENCODE: RUNX3 | $8.1 \times 10^{-16}$ | 11816 | 63 |
| | ENCODE: ELK1 | $1.5 \times 10^{-14}$ | 11349 | 61 |
| | ENCODE: YY1 | $1.5 \times 10^{-12}$ | 12289 | 177 |
| | ChEA: ETS1 | $8.0 \times 10^{-08}$ | 1359 | 20 |
| | ChEA: PADI4 | $9.4 \times 10^{-03}$ | 877 | 9 |

### Multiple outcomes

We further meta-analyzed results across all the seven breast cancer related outcomes to identify genes that are possibly associated to multiple outcomes. Since the outcomes are correlated and as a result the association p-values across the outcomes for each gene are correlated, it is difficult to use standard meta-analysis for this. In fact, the effect size estimates for association models pertaining to different types of outcomes (binary and survival), would complicate the interpretation of effect size based meta-analysis. Here, we used the cauchy combination test to meta-analyze results across the seven outcomes. 72 genes were identified to be significant at the exome-wide p-value threshold of 2.5 × 10^−06^ (Figure 3D). At FDR < 0.05, we found 508 genes to be significantly associated to the set of 7 outcomes. Although majority of these associations were driven by significant associations with at least one outcome and weaker association with several others, 13 genes were also identified (cauchy combination FDR < 0.05) which had no significant association (FDR > 0.05) to any single outcome but had possibly weaker association with multiple outcomes. For example, we found *E4F1* gene (cauchy combination test p-value = 0.028), a gene that a can induce cell growth arrest^55^, to be significant (FDR < 0.05) through the cauchy combination approach which has nominal associations with the presence/absence of lymph nodes (p-value = 0.0083), overall survival (p-value = 0.011) and HER2 status (p-value = 0.029). *DDX5* (cauchy combination test p-value = 0.021), a gene well reported to be associated to breast cancer and regulates DNA replication and cell proliferation^56^, had multiple weaker associations with the grade of tumor (p-value = 0.017), overall survival (p-value = 0.004) and HER2 status (p-value = 0.022). We conducted pathway enrichment analyses with the 508 genes that were found significant at FDR < 0.05 in meta-analyses. Similarly, as before, the results show that the numerous relevant pathways and gene-sets related to breast cancer are significantly enriched for these genes.

## Discussions

Extensive research has established that CNA are indeed important for several cancer types and subtypes, especially in breast cancers^7^. However, the intermediate mechanisms and processes via which CNA impact breast cancer related outcomes have not been conclusively established and merits further research. In this article we have outlined a novel and generalizable analytic approach to identify how CNA regulate expression levels of gene modules that ultimately influence several breast cancer related outcomes. Our approach involves two steps: using sparse canonical correlation analysis to identify gene modules associated with sets of CNA, followed by testing association between the gene modules and breast cancer related outcomes. We further carried out a meta-analysis across different types of outcomes to identify genes with multiple associations. Extensive downstream analysis shows that the genes identified through our analysis have key relevance for breast and other cancers that have also been noted in other studies. Unlike these other studies, our approach additionally identifies CNA sets that possibly regulate the genes which in turn bring about changes in outcomes related to breast cancer. Below we summarize the advantages of our analysis and its potential generalizability:

### Identifying and interpreting gene modules through sCCA

The identification of gene modules using sCCA, agnostic of the clinical or disease outcome, is a key advancement that we propose over existing work on this topic. Numerous methods have been developed to identify significant associations between individual pairs of CNA and gene expressions. However, recently several authors have hypothesized that the effects of somatic genetic variants like CNA are cascaded through complex intermediate gene network to bring about phenotypic change^57^. Thus, identifying individual CNA-gene expression associations, although informative, cannot provide deeper insight into the gene networks that can potentially be impacted by CNA. Through our joint analysis approach in sCCA, we map groups of CNA to gene modules, which essentially identifies broad gene networks potentially regulated by CNA. In fact, existing transcription factor databases show that the gene networks thus detected through sCCA have suggestive evidence to be coregulated, which indicates that such groupwise mapping approach can identify patterns of biological regulation as well.

One of the key interpretations of the gene modules is that they represent approximately independent regulatory patterns due to the orthogonality condition imposed by sCCA. Thus, in principle, the first step in our analysis, identifies key distinct biological regulatory processes that are impacted within primary breast tumor tissue. The advantage that sCCA provides over identifying single CNA-gene pairs is that, since sCCA can aggregate multiple possibly weaker association within the identified components, it can be statistically more powerful in identifying gene networks. In fact, transcriptomic studies have shown that at moderate sample size, similar to that of METABRIC study, sCCA can outperform standard pairwise regression in identifying broad downstream gene networks regulated by genetic variants. However, we acknowledge that there will possibly be a plethora of regulatory associations beyond the ones identified through sCCA, which can be identified in a larger sample.

### Association analysis using multivariable regression

The subsequent association analysis using gene modules is relatively standard and has been used commonly. However, interpreting the association p-values needs caution since the association model is a joint regression. In general, single gene vs outcome tests are different from this since the joint model additionally adjusts for correlation between the genes and reports the p-value conditional on the gene module. This multivariable regression framework identifies the genes that are associated to the clinical outcome while adjusting for the correlation within the module. While association between and outcome and a single gene can arise either due to true causality or due to correlation of the tested gene with the true causal gene, our approach accounts for the dependency and hence significant association can potentially be causal.

### Meta-analysis of results from correlated clinical outcomes

The multiple outcome meta-analysis^29^ demonstrates another advantage of our approach. In general, complex diseases like cancers, and in particular breast cancer, can have numerous biomarkers of disparate types. Combining results across the biomarkers can highlight overall important genes and genes affecting multiple biomarkers.

However, meta-analyzing association results across them is not straightforward since the results are correlated. Further for different types of biomarkers (continuous, discrete, binary, survival etc.) the effect sizes have disparate interpretation and hence standard meta-analysis might not be appropriate. However, the Cauchy combination approach alleviates these problems since it is based on p-values and not on the effect size. Further, due to the correlation agnostic property of cauchy distributions, meta-analysis of correlated p-values controls false positive rates. In principle, using this approach, meta-analysis of different types of biomarkers and outcomes are possible, as long as the univariate association mean model is correctly specified.

### Generalizable Analysis

The approach that we adopted here is highly generalizable as an overall analysis framework. The first step pertains to groupwise mapping and identification of modules followed the association analysis in the second step. Although we adopted an sCCA approach here for its ease of interpretation, several other methods mapping sets of genetic variants to gene modules can be used instead. For example, methods for biclustering and matrix factorization can be adapted in the first step to identify gene modules. In fact, groupings based on functional annotation can also be incorporated to further strengthen the mechanistic interpretation of the identified modules. The subsequent association analysis can also be customized to address scientific questions of interest. However, one of the major advantages of our pipeline is that, in principle, the two steps can be performed on separate datasets as well. For example, in current studies large scale genomic and transcriptomic data for many individuals are available while detailed information on phenotypes and traits might not be available. Thus, the identification of gene modules can be performed using such data while the subsequent association analysis can be carried out in a separate data.

Our analysis approach currently has several limitations. The optimal number of components in sCCA is chosen heuristically by maximizing the iterative ratio of canonical correlations rather than using any significance or enrichment tests. In the current set up, formulating analytical tests of significance is difficult and methods based on sCCA has mostly resorted to resampling methods. In the future, research on Bayesian formulations coupled with sequential testing is warranted to perform tests of significance for regularization and clustering methods which can indicate the optimal number of components and parameter settings.

Together, our analysis provides a comprehensive understanding of the impact of CNA can impact different breast cancer outcomes via regulation of intermediate gene networks. If a particular gene is significantly associated with a breast cancer related outcome, we can identify which set of CNA of which genomic subregion regulates it using the identified gene modules. Further, overlap of the significant genes with several breast cancer related pathways identifies the genes within the predefined biological process is differentially regulated by CNA to bring about phenotypic change. In future, as larger studies emerge with a greater coverage and spectrum of molecular phenotypes, a more comprehensive insight as to the intermediate regulatory mechanism will take shape. For that, it will be imperative to move beyond identifying single variant-gene or variant-outcome associations and conceptualize associations in context of networks and modules. The broad intuition of this analysis framework can further be extended to multi-view data sets and can be useful in integrative analysis of multi-omics data.

## Data Availability

We used clinical and genomic data publicly available at cBioPortal catalog.
Website: https://www.cbioportal.org/

## Acknowledgement

DD was supported by R01 grant from the National Human Genome Research Institute [1 R01 HG010480-01; PI Dr. Nilanjan Chatterjee]. AS and JS were supported by R01 grant from the National Cancer Institute [7 R01 CA197402-05; PI: Jaya Satagopan].

## Data and Code Availability

METABRIC: https://www.cbioportal.org/study/summary?id=brca_metabric

TCGA Breast Cancer: https://www.cbioportal.org/study/summary?id=brca_tcga

Code for analysis: https://github.com/diptavo/METABRIC_analysis

ShinyGO: http://bioinformatics.sdstate.edu/go/

FUMA: https://fuma.ctglab.nl/

CHEA: https://maayanlab.cloud/Harmonizome/dataset/CHEA+Transcription+Factor+Targets

ENCODE: https://maayanlab.cloud/Harmonizome/dataset/ENCODE+Transcription+Factor+Targets

## Notes

### Competing Interest Statement

The authors have declared no competing interest.

### Author Declarations

No new data was generated.

