## Supplementary Methods for "Sparse canonical correlation to identify breast cancer related genes regulated by copy number aberrations"

**Sample Description**

**METABRIC Study**: Molecular Taxonomy of Breast Cancer International Consortium (METABRIC) is one of the popular publicly available sources of comprehensive RNA-sequencing and copy number aberrations in breast cancer patients. The study and the overall results have been thoroughly described in Curtis et al and Pereira et al previously. In the current version of the data, 2509 patients were present overall with detailed sample characteristics, recorded clinical outcomes and cancer biomarkers. Among these 1904 patients had gene-expression data on 24,360 genes across the genome in primary breast tumors. Further, these patients also had data on 22544 somatic copy number aberrations (CNA). Each of the CNA were physically mapped to a proximal gene. We used data on CNA, gene expression and clinical outcomes for the 1904 who had complete data.

**TCGA Breast Invasive Carcinoma**: This is a legacy data set previously known as TCGA Provisional. The data was gathered as part of the Broad Institute of MIT and Harvard Firehose initiative, a cancer analysis pipeline. The dataset contains clinical and genomic data for 1,108 carcinomas for 1,101 patients. From this we used the complete genomic data of 1,077 patients, across 24,777 CNA sites and 20,441 gene expressions. Similar to METABRIC data, each CNA were physically mapped to a proximal gene.

**Extended Methods**

To introduce sparse canonical correlation for identifying gene modules, we first describe standard canonical correlation (CC) analysis. For *n* individuals, let $\boldsymbol{G}^{\boldsymbol{n\times p}}$ be the normalized matrix for *p* CNA sites and $\boldsymbol{E}^{\boldsymbol{n\times g}}$ be the normalized gene-expression matrix for *g* genes all of which are distant (*trans*) to the variants. The classical CC analysis seeks to find linear combinations of CNA ($\boldsymbol{u}^{\boldsymbol{p\times1}}$) and gene expressions ($\boldsymbol{v}^{\boldsymbol{g\times1}}$) such that the correlation between $\boldsymbol{Gu}$ and $\boldsymbol{Ev}$ is maximized i.e.,

$$\left( \boldsymbol{u,v} \right)\boldsymbol{=}\boldsymbol{argmax}{\tilde{\boldsymbol{v}}}^{\boldsymbol{T}}{\tilde{\boldsymbol{E}}}^{\boldsymbol{T}}\tilde{\boldsymbol{G}}\tilde{\boldsymbol{u}}$$

with $\boldsymbol{||}\tilde{\boldsymbol{u}}\boldsymbol{||}_{\boldsymbol{2}}\boldsymbol{= 1}$ and $\boldsymbol{||}\tilde{\boldsymbol{v}}\boldsymbol{||}_{\boldsymbol{2}}\boldsymbol{= 1}$

where $\boldsymbol{||.}\boldsymbol{||}_{\boldsymbol{h}}$denotes the *L_h_* norm. The subsequent pairs of CC components are obtained by similarly maximizing the correlation between $\tilde{\boldsymbol{G}}\boldsymbol{u}$ and $\tilde{\boldsymbol{E}}\boldsymbol{v}$ and under the constraint of being uncorrelated or orthogonal to the previous components. In practice, this can be obtained from the spectral decomposition of $\boldsymbol{W}^{\boldsymbol{T}}\boldsymbol{W}$ where ${\boldsymbol{W}= \tilde{\boldsymbol{E}}}^{\boldsymbol{T}}\tilde{\boldsymbol{G}}$. If $\left( \boldsymbol{u}_{\boldsymbol{k}}\boldsymbol{,}\boldsymbol{v}_{\boldsymbol{k}} \right)$ denote the $\boldsymbol{k}^{\boldsymbol{th}}$ pair of CC components, then

$\boldsymbol{u}_{\boldsymbol{k}}\boldsymbol{=}\boldsymbol{k}^{\boldsymbol{th}}$ eigen-vector of $\boldsymbol{W}^{\boldsymbol{T}}\boldsymbol{W}$

and $\boldsymbol{v}_{\boldsymbol{k}}\boldsymbol{=W}$

The CC components can be interpreted as linear latent linear factors which explain the correlation (or covariance, if not normalized) between the CNA and the gene-expressions by aggregating multiple, possibly weaker, associations. However, it is difficult to interpret the CC components directly since all CNA and gene-expressions can have non-zero coefficients in the $\left( \boldsymbol{u}_{\boldsymbol{k}}\boldsymbol{,}\boldsymbol{v}_{\boldsymbol{k}} \right)$.

**Sparse canonical correlation (sCCA)**

For better interpretation, we proposed to derive CC components that will involve only limited number of CNA and genes based on sCCA. Thus, we introduce penalty terms to regularize the CC components, effectively reducing the smaller components to 0 for a suitable penalty. We use an L_1_ penalty, equivalent to LASSO. The optimization problem is given by

$$\left( \boldsymbol{u,v} \right)\boldsymbol{=}\boldsymbol{argmax}{\tilde{\boldsymbol{v}}}^{\boldsymbol{T}}\boldsymbol{W}\tilde{\boldsymbol{u}}$$

with $\boldsymbol{||}\tilde{\boldsymbol{u}}\boldsymbol{||}_{\boldsymbol{1}}\boldsymbol{\leq}\boldsymbol{c}_{\boldsymbol{u}}$ ;$\boldsymbol{||}\tilde{\boldsymbol{v}}\boldsymbol{||}_{\boldsymbol{1}}\boldsymbol{\leq}\boldsymbol{c}_{\boldsymbol{v}}$and $\boldsymbol{||}\tilde{\boldsymbol{u}}\boldsymbol{||}_{\boldsymbol{2}}\boldsymbol{= 1}$, $\boldsymbol{||}\tilde{\boldsymbol{v}}\boldsymbol{||}_{\boldsymbol{2}}\boldsymbol{= 1}$

L_1_ penalty produces sparse solution and thus allows for variable selection and, hence, more effective downstream interpretation. The sCCA components $\left( \boldsymbol{u,v} \right)$ denote the loadings of the CNA and genes respectively. A non-zero element in $\boldsymbol{u}$ (or $\boldsymbol{v}$) implies that the correlation between linear combinations of the respective CAN and gene is maximized.

Here we have not corrected for the correlation between CNA or gene-expressions, i.e., the column correlations of $\tilde{G}$ or $\tilde{E}$. This can lead to selection of highly correlated CNA (or gene expressions) as a group through the non-zero elements of $u$ (or $v$). This approach is known to have superior performance compared to canonical correlations based on correlation correction, and can lead to detection of coregulation and interpretable dependency structures between the CNA and gene-expressions (Witten et al., YEAR).

For a fixed $\boldsymbol{u=}\boldsymbol{u}_{\boldsymbol{0}}$, the objective function is a standard LASSO optimization problem in $v$:

$\boldsymbol{v=}\boldsymbol{argmax}{\tilde{\boldsymbol{v}}}^{\boldsymbol{T}}\boldsymbol{W}\boldsymbol{u}_{\boldsymbol{0}}$ with $\boldsymbol{||}\tilde{\boldsymbol{v}}\boldsymbol{||}_{\boldsymbol{1}}\boldsymbol{\leq}\boldsymbol{c}_{\boldsymbol{v}}$ and $\boldsymbol{||}\tilde{\boldsymbol{v}}\boldsymbol{||}_{\boldsymbol{2}}\boldsymbol{= 1}$

and similarly, for a fixed $uv\boldsymbol{=}{vu}_{\boldsymbol{0}}$

$\boldsymbol{u=}\boldsymbol{argmax}\boldsymbol{v}_{\boldsymbol{0}}^{\boldsymbol{T}}\boldsymbol{W}\tilde{\boldsymbol{u}}$ with $\boldsymbol{||}\tilde{\boldsymbol{u}}\boldsymbol{||}_{\boldsymbol{1}}\boldsymbol{\leq}\boldsymbol{c}_{\boldsymbol{u}}$ and $\boldsymbol{||}\tilde{\boldsymbol{u}}\boldsymbol{||}_{\boldsymbol{2}}\boldsymbol{= 1}$

Hence, this optimization problem can be solved iteratively (Witten et al., YEAR).

$c_{u}$ $c_{v}$

The canonical correlation value is defined as

$$q\boldsymbol{=}\boldsymbol{v}^{\boldsymbol{T}}\boldsymbol{Wu}$$

Given the k^th^ of sCCA components $\left( \boldsymbol{u}_{\boldsymbol{k}}\boldsymbol{,}\boldsymbol{v}_{\boldsymbol{k}} \right)$, we use Hotelling’s deflation with the above algorithm to extract the (k+1)^th^ sCCA component as the leading eigen vector of:

$$\boldsymbol{W}_{\boldsymbol{k+1}}\boldsymbol{=}\boldsymbol{W}_{\boldsymbol{k}}\boldsymbol{-|q|}\boldsymbol{v}_{\boldsymbol{k}}{\boldsymbol{u}_{\boldsymbol{k}}}^{\boldsymbol{T}}$$

$$\boldsymbol{W}_{\boldsymbol{1}}=\boldsymbol{W}$$

**Choice of sparsity parameters.** Most applications of variable selection use cross-validation techniques to determine the choice the tuning parameters$c_{v}$and $c_{u}$that will maximize the prediction accuracy in an independent test sample. Our pivotal goal is interpretation of sCCA components when the numbers of CNA and gene expressions are much higher in dimension compared to the sample size. Therefore, we used an intersection-minimization approach to improve the interpretation of the sCCA components. Two sCCA components $u_{j}$ and $u_{k}$ are orthogonal in CNA if the two components do not have the same elements with non-zero loadings. Equivalently, the intersection of non-zero elements of the two components is a null set. However, in sCCA orthogonality cannot be guaranteed by the estimation algorithm, although the resulting components are approximately orthogonal. In our estimation, we aim to minimize the intersection of the CNA sites selected in successive CNA components to ensure that we capture independent patterns of coregulation. We chose the sparsity parameter $\boldsymbol{c}_{\boldsymbol{u}}$ such that the none of the extracted CNA components had any common CNA sites selected.

We aimed to choose $\boldsymbol{c}_{\boldsymbol{v}}$ to achieve relatively small set of selected genes. We imposed $\boldsymbol{||}\tilde{\boldsymbol{v}}\boldsymbol{||}_{\boldsymbol{0}} \leq100$ for all the gene-components, meaning fewer than 100 genes would be selected in each component i.e., fewer than 100 genes would have non-zero values in each gene component $v$. Given the $\boldsymbol{c}_{\boldsymbol{u}}$, we then obtained $\boldsymbol{c}_{\boldsymbol{v}}$ through a standard grid search which maximized the canonical correlation $q$ under the above constraint.

${GS}_{i}$ ${CS}_{i}$ ${GS}_{i}$ ${CS}_{i}.$ ${CS}_{-i}$ ${CS}_{-i}$ ${CS}_{-i}|= {|CS}_{i}|$ ${CS}_{-i}$ ${CS}_{i}$ ${GS}_{i}$ ${CS}_{-i}$ ${GS}_{i}$ ${GS}_{i}$ ${CS}_{i}$ ${GS}_{i}$ ${CS}_{i}$ ${GS}_{i}$ ${CS}_{i}$ ${GS}_{i}$ ${CS}_{i}$ ${GS}_{i}$ ${CS}_{i}$ ${GS}_{i}$ ${CS}_{-i}$

**Resampling test for TCGA data.** We validated the CAN and gene expressions selected via sCCA from METABRIC data using the TCGA breast cancer data. The goal of this validation was to establish that the CNA and gene expressions selected from METABRIC also have statistically significant correlations in TCGA. We developed the following resampling approach for validation.

Denote $\hat{u}_{1}, \hat{u}_{2}, \ldots, \hat{u}_{C}$, denote C CNA components and $\hat{v}_{1}, \hat{v}_{2}, \ldots, \hat{v}_{E}$ denote E gene expression components identified via sCCA in METABRIC data. Let $p_{i}^{C}$ and $g_{i}^{E}$ denote the numbers of non-zero elements of the i-th components $\hat{u}_{i}$ and $\hat{v}_{i}$, respectively. Equivalently, $p_{i}^{C}$ CNAs and $g_{i}^{E}$ gene expressions were selected by the i-th components in the analysis of METABRIC data. Denote $G_{i}^{(TCGA)}$ and $E_{i}^{(TCGA)}$denote the $n\times p_{i}^{C}$ matrix $n\times g_{i}^{E}$ matrix of these CNAs and gene expressions, respectively, for the n TCGA subjects. Calculate the squared correlation between every column of $G_{i}^{(TCGA)}$ and every column of $E_{i}^{(TCGA)}$. Let $\rho_{i}^{2}$ denote the average of these $p_{i}^{C}\times g_{i}^{E}$ squared correlations.

To determine whether the average squared correlation between these $p_{i}^{C}$ CNAs and $g_{i}^{E}$ gene expressions corresponding to the i-th sCCA component is significantly different from 0, we use the following resampling approach. Keeping the matrix of the selected CNAs fixed, i.e., keeping $G_{i}^{(TCGA)}$ fixed, randomly select $g_{i}^{E}$ gene expressions and obtain $E_{i}^{(TCGA, rand)}$ as the corresponding $n\times g_{i}^{E}$ matrix of gene expressions. Obtain $r_{i}^{2}$ as the average of the squared correlation between the columns of $G_{i}^{(TCGA)}$ and $E_{i}^{(TCGA, rand)}$. Repeat this procedure B times – for example, B = 1,000 – to obtain the conditional null distribution of the squared correlation across gene expression for fixed CNAs in $G_{i}^{(TCGA)}$. The p-value is the fraction of the resulting $r_{i}^{2}$that exceed $\rho_{i}^{2}$. We term this as gene component validation. Similarly, we can fix the gene expressions $E_{i}^{(TCGA)}$ and evaluate the statistical significance of its average squared correlation with the $p_{i}^{C}$ CNAs to obtain CNA validation in the same vein.

This is a valid replication since the TCGA and METABRIC data do not overlap in terms of samples. Further, we evaluate the average squared correlations between CNAs and gene expressions within the TCGA data. This ensures comparability of the values while controlling for internal validity. Our results show that most of the gene components and CNA components are strongly validated (p-value < 0.001) while several are weakly validated (0.001 < p-value < 0.05). This indicates that the CNAs and gene expressions selected in METABRIC data have evidence of strong association externally as well and indicate that systematic biases in selection might not be present.

Comparison of model fitting in TCGA data: To ascertain the relevance of the genes identified to be significantly associated to ER status, we used a model fitting comparison in TCGA data. As a benchmark, we used a sparse logistic regression in METABRIC data to identify gene expressions that are significantly associated with ER status. The difference between this and the sCCA-based approach is that this sparse logistic regression is completely agnostic of regulatory CNA and relies on predictive performance metrics to select genes. Thus, sparse logistic regression considers the full set of 24,360 genes whereas our sCCA-based approach is restricted to only 831 genes across 14 components. In METABRIC data, the sparse logistic regression selects a set of 161 genes. Thus, we have two candidate sets of genes identified through (1) sCCA-based approach (2) sparse logistic regression.

Next, using the TCGA data and the genes selected in METABRIC data, we fit two logistic model in the TCGA data using the genes identified through (1) sCCA-based approach (2) sparse logistic regression and compare the BIC for each model. The BIC is a measure of the goodness of fit using the respective set of covariates. If a particular set of covariates is a substantially better fit for the outcome, the corresponding BIC is expected to be smaller in comparison to other models. In general, not all genes in METABRIC are present in TCGA although there is substantial intersection. We use the genes common between TCGA and METABRIC for this analysis.

**Supplementary Figure:**

S1: Results for validation study of genes and CNA identified through sCCA on METABRIC data in TCGA Firehose legacy data.

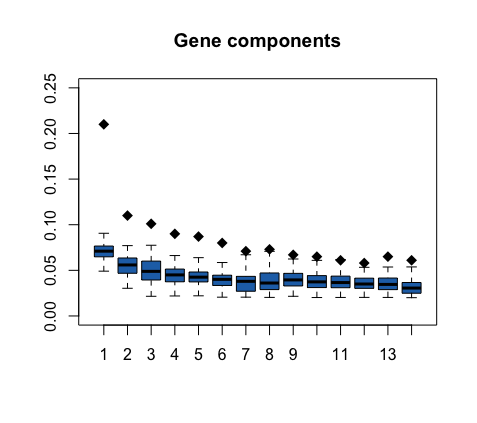

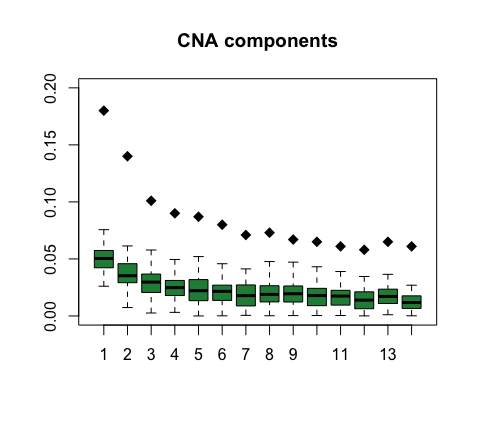
